# Manufacturing Processes of Peanut (*Arachis hypogaea*) Allergen Powder-dnfp

**DOI:** 10.1101/2022.06.28.22276947

**Authors:** Stephanie A. Leonard, Yasushi Ogawa, Paul T. Jedrzejewski, Soheila J. Maleki, Martin D. Chapman, Stephen A. Tilles, George Du Toit, S. Shahzad Mustafa, Brian P. Vickery

## Abstract

Important components of drug safety, efficacy, and acceptability involve manufacturing and testing of the drug substance and drug product. Peanut flour sourcing/processing and manufacturing processes may affect final drug product allergen potency and contamination level, possibly impacting drug safety, quality, and efficacy. We describe key steps in the manufacturing processes of peanut (*Arachis hypogaea*) allergen powder-dnfp (PTAH; Palforzia®), a drug used in oral immunotherapy (OIT) for the treatment of peanut allergy. Established criteria for source material must be met for manufacturing PTAH drug product. Degree of roasting was determined with a Hunter colorimeter. Protein/allergen content, identity, potency, safety, and quality of each batch of PTAH drug substance were assessed with a combustion analyzer, allergen-specific Western blot (immunoblotting), ELISA, and HPLC; contaminants (i.e., aflatoxin) were measured by UPLC. Roasting degree beyond “light roast” was associated with variable degrees of protein allergen degradation, or potentially aggregation, particularly for Ara h 1 and Ara h 3. Relative potency and amounts of protein allergens showed variability due in part to seasonal/manufacturing variability. Proportion of lots not meeting aflatoxin limits has increased in recent years. Up to 60% of peanut flour source material failed to meet screening selection acceptance criteria for proceeding to drug substance testing, mostly because of failure to meet potency acceptance criteria. Other lots were rejected due to safety and quality. Influence of potency variation, within specification parameters, on safety/tolerability observed in trials was considered low, in part due to stringent controls placed at each step of manufacture. Extensive variability in allergen potency is a critical issue during immunotherapy, particularly during OIT initial dose escalation and up-dosing, as it may result in lack of efficacy or avoidable adverse allergic reactions. Based on EU and US regulatory requirements, the production of PTAH includes manufacturing controls to ensure drug product safety, potency, and quality. For example, although PTAH contains all peanut allergens, each lot has met strict criteria ensuring consistent allergenic potency of Ara h 1, Ara h 2, and Ara h 6. The rigor of PTAH’s manufacturing process ensures reliable dose consistency and stability throughout its shelf life.

## 1 Introduction

A critical component of drug safety, efficacy, and acceptability involves controls across the manufacturing and testing process of a drug substance and drug product (1, 2). A drug substance is an active ingredient intended to provide acceptable pharmacological activity or other direct effect used in the diagnosis, cure, mitigation, treatment, or prevention of disease or to affect the structure or function of the human body (3). A drug product is the finished dosage form that contains the drug substance and may include other active or inactive ingredients (3). In the case of oral immunotherapy (OIT) for food allergies, the use of food as the allergen source is intended to have medical and therapeutic effects, as opposed to food that is intended as nutrition (4, 5).

In recent decades, the application of the concepts of “Good Manufacturing Practice” (GMP) to “allergen standardization” has emerged as key regulatory priority and the United States (US) Food and Drug Administration (FDA) has issued multiple “Guidance for Industry” documents outlining Chemistry Manufacturing and Controls Guidance for allergens used for diagnosis or treatment (1, 6). In Europe, similar regulatory guidelines exist (7). GMP refers to standards of production, including the physical facilities, equipment, personnel, and other processes involved with allergen manufacturing (1). Despite adherence to GMP, allergens are highly heterogenous, partly because they are derived from natural sources, but also because manufacturing may involve roasting, grinding, defatting, extraction, clarification, and sterilization (8-11) that may introduce allergen heterogeneity.

Allergen standardization refers to maintaining consistency within manufacturing process and analytical capabilities between lots of allergen products and between products from different manufacturers; it is intended to improve both safety and efficacy of allergen immunotherapy (12). This requires the use of rigorously qualified and highly characterized reference standards against which each lot must be measured for potency (i.e., allergenic activity) as well as other quality attributes (e.g., identity). Although not all allergens used in immunotherapy are standardized, for allergens compounded and administered as immunotherapy by practicing allergists in the US (primarily inhalant or venom allergens via subcutaneous injection), the Joint Task Force on Practice Parameters, representing the American Academy of Allergy, Asthma, and Immunology (AAAAI) and the American College of Allergy, Asthma, and Immunology (ACAAI), practice parameter on immunotherapy recommends using standardized allergens when available:

“…standardized extracts should be used to prepare allergen immunotherapy treatment sets…The advantage of standardized extracts is that the biologic activity is more consistent, and therefore the risk of an adverse reaction caused by extract potency variability should be diminished.” (8)

Similarly, the European Academy of Allergy and Clinical Immunology (EAACI) guidelines on allergen immunotherapy acknowledge the “…need to limit practice to the use of high-quality, standardized allergen immunotherapy products with good evidence of effectiveness…as many available products are not supported by sufficient evidence of efficacy” (13). Additionally, several publications have been prepared by the EAACI Taskforce on Regulatory Aspects of Allergen Immunotherapy and are part of the EAACI Allergen Immunotherapy Guidelines (2, 14). Comparisons of allergen manufacturing and quality control regulations between the US and European Union (EU) have been reviewed previously (14, 15).

In the early 2000s, OIT emerged as a promising strategy based on small, placebo-controlled studies at academic centers and small, uncontrolled studies conducted by private practitioners (16-18). In 2011, a research retreat was organized and sponsored by an advocacy group called the Food Allergy Initiative (now known as Food Allergy Research and Education) (19). This retreat included a variety of stakeholders, including patient advocates, clinicians, pharmaceutical industry members, and representatives from both the National Institutes of Health and FDA. A consensus was reached that there was a significant unmet need for a standardized OIT approach to food allergy treatment. This led to formation of the Allergen Research Corporation (later renamed Aimmune Therapeutics) (20).

After completing both phase 2 and phase 3 clinical trials (21-23), peanut (*Arachis hypogaea*) allergen powder-dnfp (PTAH; Palforzia®) was approved in 2020 by both the US FDA and the EU European Commission (24, 25). “dnfp” refers to the four-letter suffix extension assigned to PTAH and a naming convention applying to biological products as required by the FDA (26). In the US, PTAH is indicated for the mitigation of allergic reactions, including anaphylaxis, that may occur with accidental exposure to peanut (24, 25). PTAH is approved in the US and EU for use in patients with peanut allergy aged 4 through 17 years and is administered using a standard escalating-dose program (24, 25).

Source material used for PTAH is a naturally produced material subject to a manufacturing process and storage conditions that impact its use as an approved pharmaceutical product (27, 28). An overview of the multiple quality assurance steps (ie, unit operations and process controls) associated with the manufacture of source material is shown in **Figure 1**. The source material for PTAH is 12% defatted, lightly roasted peanut flour produced by the Golden Peanut and Tree Nuts (GPTN) company, and GPTN independently tests for quality and safety attributes in compliance with food GMP requirements.

**Figure 1.**
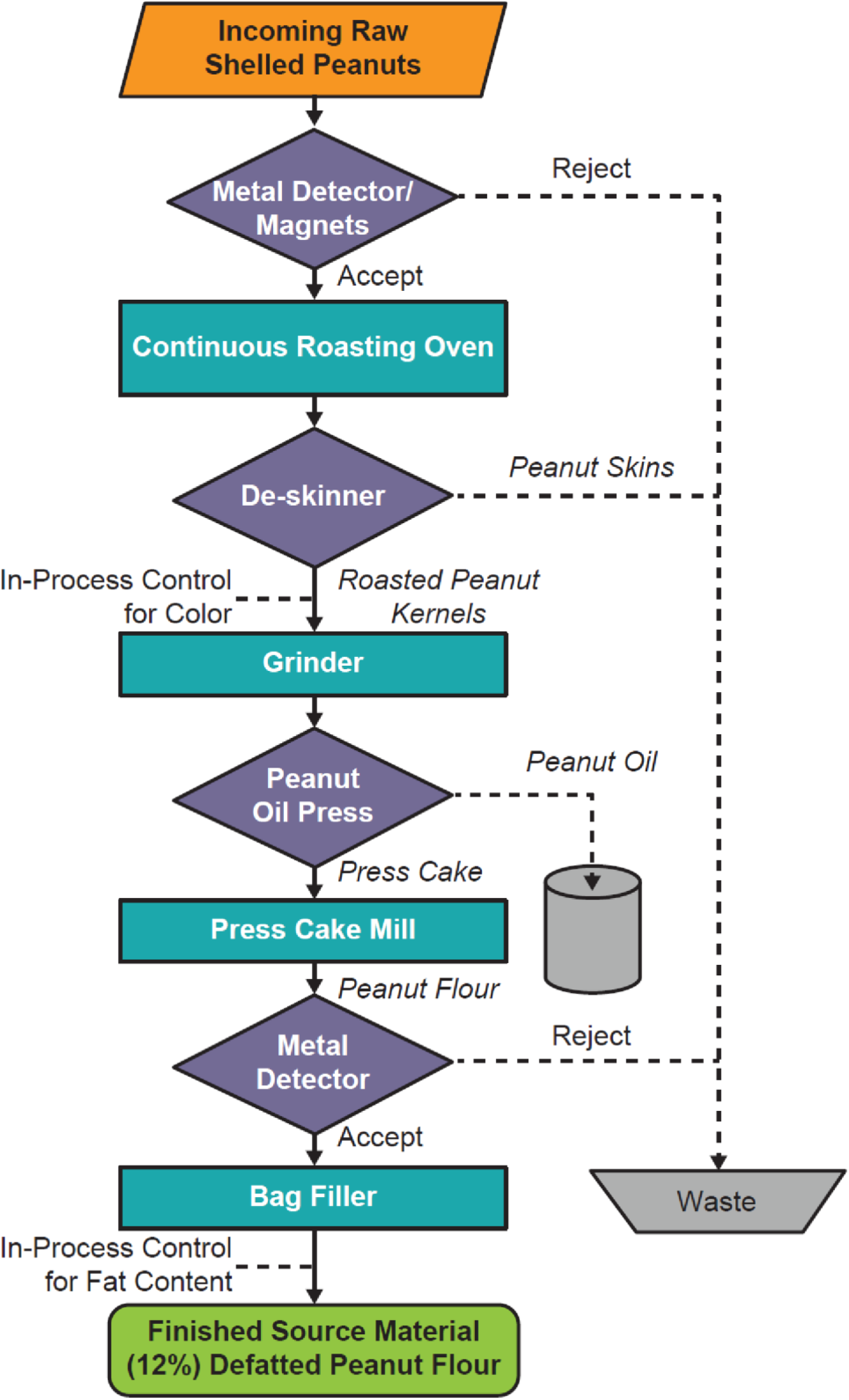
Flowchart of the Manufacturing Process for Peanut Flour Source Material Generation at GPTN. **Abbreviation:** GPTN, Golden Peanut and Tree Nuts.

These raw peanuts conform to the Code of Federal Regulations Title 7 Part 996, Minimum Quality and Handling Standards for Domestic and Imported Peanuts Marketed in the US (ie, for consumption as food), which limits content of damaged kernels and ensures the peanut stock is minimized for *Aspergillus flavus*, the fungus responsible for aflatoxin (a poisonous carcinogen) contamination in crops (29). The allowable limit of total aflatoxin level in peanut flour distributed as food is 15 parts per billion (ppb) (29). To allow for variations incident to proper grading and handling, a tolerance by weight of 5% split peanut kernels is allowed. Split kernels, due to approximately 50% more surface area per unit mass than the intact kernels, would be exposed to more heat than intact kernels during the roasting process, which can affect allergen quality.

This manuscript will describe the manufacturing of PTAH, the first US FDA and EU European Commission–approved OIT, from the source material of peanut flour to drug substance to the final drug product. We explain the process of peanut source material selection and processing prior to drug substance testing and report the testing and standards for transforming peanut flour material into drug substance.

## 2 Methods

### 2.1 Manufacturing of Palforzia: from source material to drug substance/drug product

To ensure peanut flour source material batches designated as drug substance can be used to manufacture PTAH drug product of consistent safety and quality, the source material batches are subject to a selection process before undergoing formal testing and release as drug substance into GMP production. After receipt and sampling at the testing facility, PTAH drug substance in its container (high-density polyethylene-lined paper bags placed inside a secondary container closure system, a high-density polyethylene drum lined with two low-density polyethylene bags for protection) is stored at 2ºC to 8ºC. Stability is monitored for at least 36 months to ensure lots remain stable within their shelf lives. The stability monitoring of the final drug product (in capsules and blister packed or in sachets) is also conducted up to 48 months to ensure the potency, safety, and quality of the product remain within specification until the end of its shelf life.

### 2.2 Source material

The source material selection process for PTAH is rigorous (**Figure 2**); all clinical study lots are manufactured from 12% fat, lightly roasted peanut flour from GPTN. Source material is stored in the warehouse under ambient conditions until it is shipped for testing and released as drug substance and manufacture into drug product under pharmaceutical GMPs. The first step in the source material selection process is an evaluation of the results reported on the GPTN certificates of analysis (COA) for the batches being considered. Results on the COA for the source material batch are also evaluated for alignment with acceptance criteria for PTAH drug substance specification for microbiological quality attributes and aflatoxins, as these criteria are more stringent than for GPTN source material.

**Figure 2.**
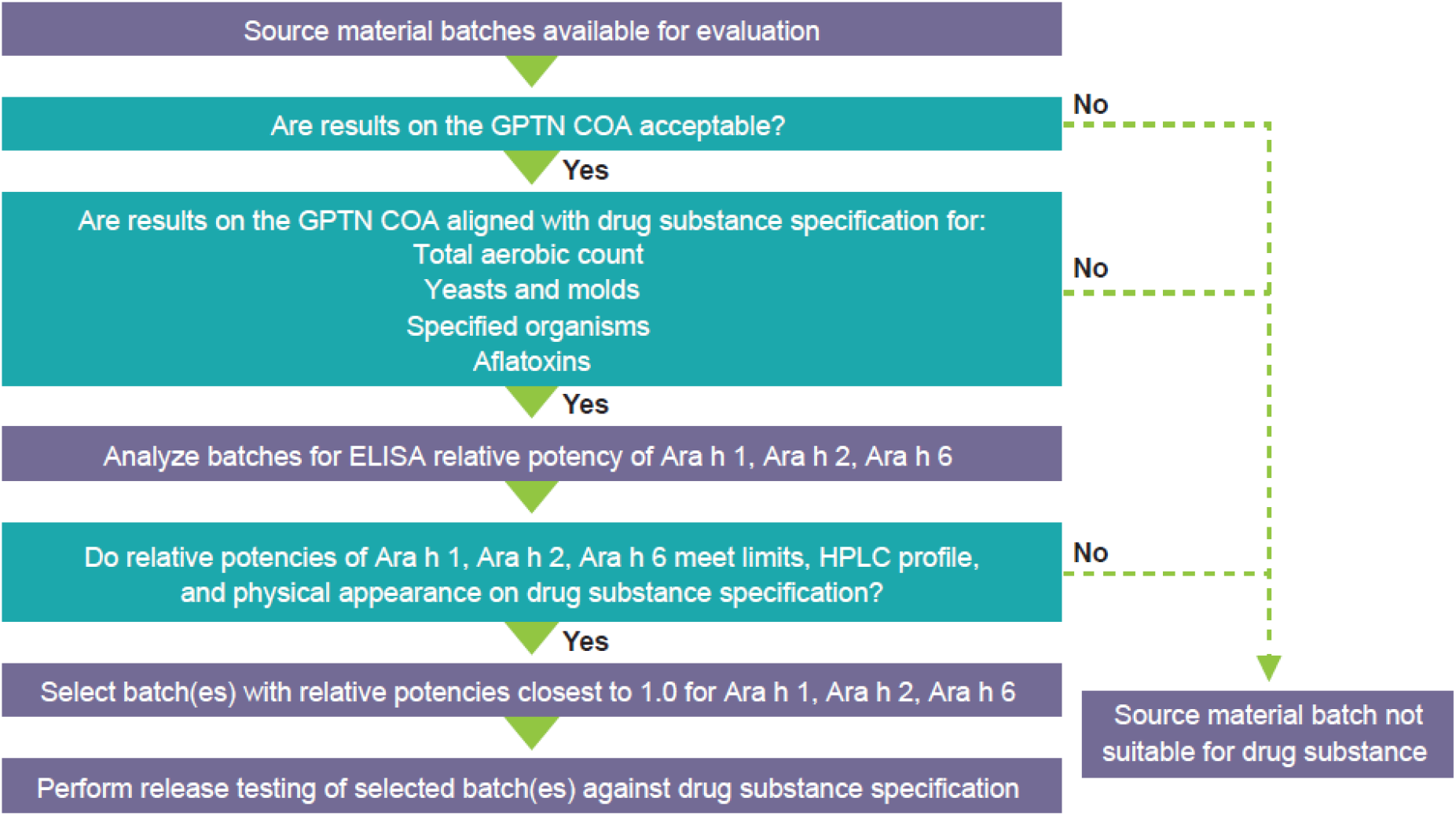
PTAH Source Material Selection Process. **Abbreviations:** COA, Certificate of Analysis; ELISA, enzyme-linked immunosorbent assay; GPTN, Golden Peanut and Tree Nuts; HPLC, high-performance liquid chromatography; PTAH, peanut (*Arachis hypogaea*) allergen powder-dnfp.

### 2.3 Evaluation of bulk peanut flour lots

Evaluation of source material peanut flour lots for pre-selection involves testing for 1) total protein content and quality and relative potency of each of three immunodominant allergens, Ara h 1, Ara h 2, and Ara h 6; 2) aflatoxins, including aflatoxin B_1_ and total aflatoxin; 3) high-performance liquid chromatography (HPLC) relative percent area profile of extracted proteins; and 4) physical appearance, including color, texture, and inherent attributes. Protein content was measured by nitrogen content determined by a Dumatherm nitrogen/protein analyzer (Gerhardt, Königswinter, Germany) using protein/nitrogen conversion factor 5.46. Allergen-specific antibodies custom prepared were used in Western blot and enzyme-linked immunosorbent assay (ELISA) (Aimmune developed) methods, validated using internal reference standards. To determine if allergens were intact and confirm immunoassays, product-specific (Aimmune developed) HPLC analysis was used. Briefly, the method used involved a C18 column resin and gradient for elution with ultraviolet detection. The methods have been developed and validated to ensure consistency and robustness in global laboratories. The proportion of screened lots rejected as unsuitable for drug substance was reported.

In the absence of a reference standard specified by regulatory authorities for peanut, the PTAH manufacturing process uses internal reference standards (28). Of note, allergens in currently marketed products are from natural allergen sources and standardization of these products is generally based on internal references and assays (15). These reference standards were prepared from a selected lot of peanut flour that was extensively tested and characterized to establish its potency (27, 28), allergen profile, and quality. The primary reference standard is assigned a nominal potency value of 1.0, stored long term, and is used to qualify secondary reference standards, which, in turn, are used in routine lot testing. Additional detail on the primary and secondary reference standards is found in the **Supplementary Material**.

**Table 1** lists attributes for selected screening tests applied to lots of source material peanut flour being considered for drug substance. In addition to controlling for Ara h 1, Ara h 2, and Ara h 6, other allergens including Ara h 3, Ara h 7, Ara h 8, Ara h 9, and Ara h 10 through Ara h 17, which may be considered less clinically relevant yet may be predictive for outcomes such as systemic allergic reactions or epinephrine use or discontinuation due to gastrointestinal adverse events, have also been shown to bind immunoglobulin E. These tests include measurements of relative levels of allergens (by HPLC) and relative potency of immunodominant allergens compared with an internally qualified reference standard using ELISA, protein integrity by HPLC, and levels of aflatoxins using ultra-performance liquid chromatography (UPLC); color, fat content, moisture, and microbes were analyzed by compendial methods.

**Table 1.** Selected Tests Used During Peanut Flour Source Material Screening.

| Test | Attribute |
| --- | --- |
| Relative potency by ELISA<sup>a</sup> | Ara h 1 |
|  | Ara h 2 |
|  | Ara h 6 |
| Protein integrity and content by HPLC | Area % Ara h 2 |
|  | Area % Ara h 6 |
| Aflatoxin by UPLC | Aflatoxin B<sub>1</sub> |
|  | Total aflatoxins |
| Color | L scale |
| Fat content | % by weight |
| Moisture | % by weight |
| Aerobic plate count | CFU/g |
| Yeast and mold count | CFU/g |

<sup>a</sup>Relative potencies of the peanut allergens are determined by testing against a peanut flour reference standard, which has assigned potency of 1.0 for each allergen. Protein integrity HPLC profile is assessed against the profile of peanut flour reference standard and must be qualitatively comparable. In addition, % peak area of Ara h 2 and Ara h 6 allergens relative to the total peak areas are reported (**Figure 4**).
**Abbreviations:** CFU, colony-forming unit; ELISA, enzyme-linked immunosorbent assay; HPLC, high-performance liquid chromatography; UPLC, ultra-performance liquid chromatography.

### 2.3 End points and assessments

The identity, potency, and purity of each batch of PTAH drug substance were assessed and confirmed according to the specifications, in accordance with the International Conference of Harmonization (ICH) Q6A and Q6B. Source material batches accepted for formal testing as PTAH drug substance were tested according to the drug substance release specification and released into GMP production of drug product.

We report findings related to allergen content, aflatoxin, and bioburden of PTAH of source material received from GPTN. We also report the proportion of screened lots rejected as unsuitable for drug substance testing from the years 2018 to 2021 and descriptively compare clinical findings from clinical trials. Correlation and/or associations of relative potency to clinical outcomes from previously published phase 3 clinical trials of PTAH are also reported (22, 23).

## 3 Results

Established critical limits and in-process controls for manufacturing the source material (**Table 2**) must be met to ensure the suitability of peanut flour for drug substance screening and its use in further manufacturing of the allergen source material into PTAH drug product. Additional attributes verified by GPTN (or its contract test laboratories) and documented in the COA include protein content, fat content, moisture, ash, color, aerobic plate count, yeast and mold count, coliform count, *E. coli* count, *E. coli* O157:H7, *Salmonella, L. monocytogenes*, and *S. aureus*.

**Table 2.**
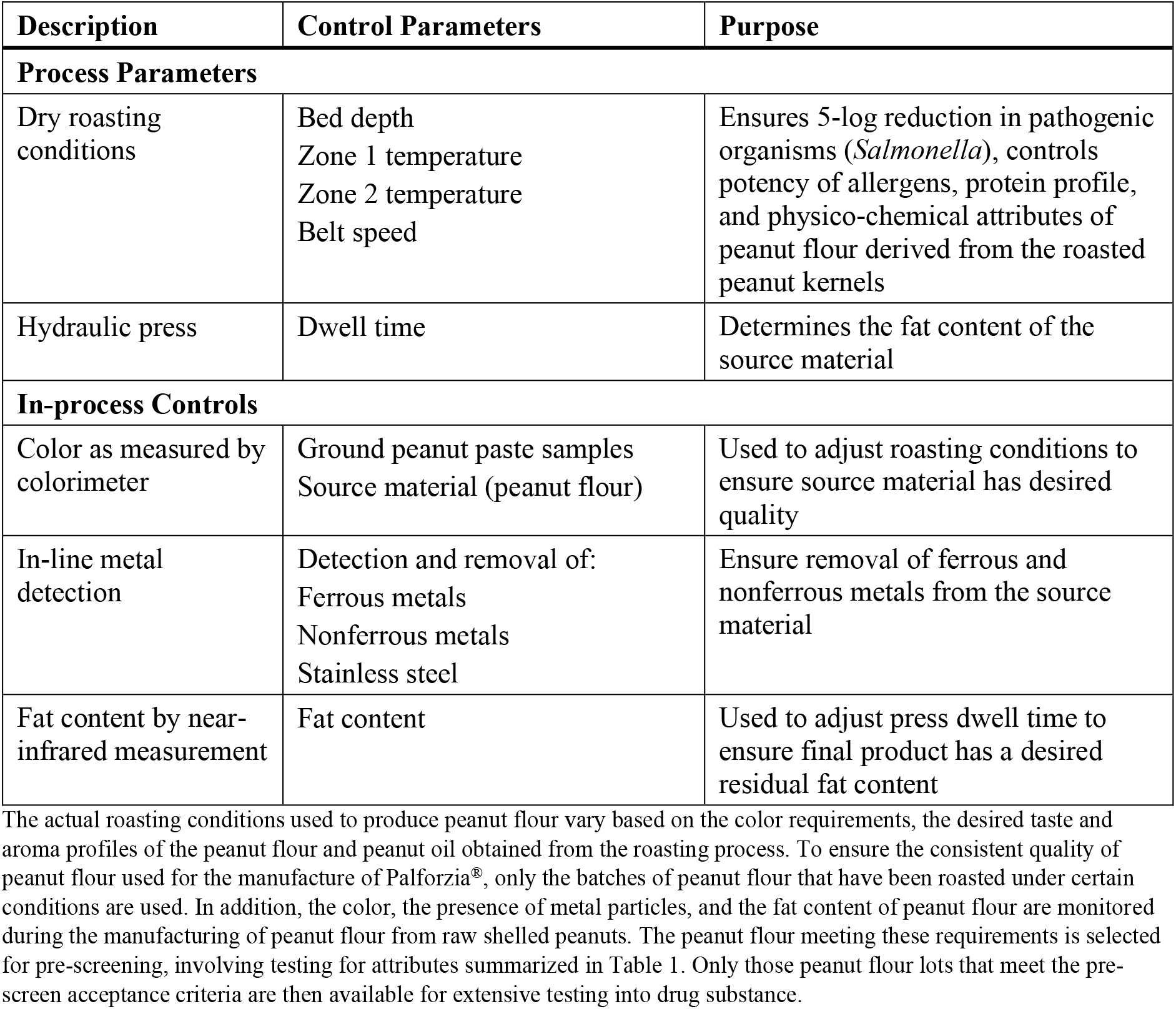
Manufacturing Process Parameters and In-Process Controls for the Allergen Source Material.

| Description | Control Parameters | Purpose |
| --- | --- | --- |
| **Process Parameters** |  |  |
| Dry roasting conditions | Bed depth<br>Zone 1 temperature<br>Zone 2 temperature<br>Belt speed | Ensures 5-log reduction in pathogenic organisms (*Salmonella*), controls potency of allergens, protein profile, and physico-chemical attributes of peanut flour derived from the roasted peanut kernels |
| Hydraulic press | Dwell time | Determines the fat content of the source material |
| **In-process Controls** |  |  |
| Color as measured by colorimeter | Ground peanut paste samples<br>Source material (peanut flour) | Used to adjust roasting conditions to ensure source material has desired quality |
| In-line metal detection | Detection and removal of:<br>Ferrous metals<br>Nonferrous metals<br>Stainless steel | Ensure removal of ferrous and nonferrous metals from the source material |
| Fat content by near-infrared measurement | Fat content | Used to adjust press dwell time to ensure final product has a desired residual fat content |

### 3.1 Degree of roasting and color

The impact of degree of roasting on the peanut source material is demonstrated using HPLC testing by peak area proportions of the peanut allergens Ara h 2, Ara h 6, Ara h 1, and Ara h 3 in the elution profiles and is shown in **Table 3**. The peak areas are tabulated relative to the total peak area of the allergen in light roasted peanut flour. These results suggest that roasting beyond “light roast” affects allergen content, which is shown to be more pronounced for Ara h 1 and Ara h 3. In addition, the degree of roasting affects the color (data not shown) of the peanut source material as follows: the light roasted being less roasted while the dark roast receives more of the roasting conditions (temperature and time). The roasting process imparts significant chemical processes to the protein allergens (i.e., glycation through Maillard reaction; protein crosslinking through inter- and intraprotein bonding changes).

**Table 3.** Source Material Allergens Content in 12% Fat Peanut Flour Determined by HPLC in Peanut Flour Manufactured Under Variable Degrees of Roasting Conditions.

| Type of Source Material | % Area of Major Allergens<sup>a</sup> |  |  |  |
| --- | --- | --- | --- | --- |
|  | Ara h 2 | Ara h 6 | Ara h 1 | Ara h 3 |
| 12% fat peanut flour, light roast (Runner type) | 100 | 100 | 100 | 100 |
| 12% fat peanut flour, medium roast (Virginia type) | 66.5 | 53.9 | 59.0<sup>b</sup> | 35.6 |
| 12% fat peanut flour, dark roast (Runner type) | 77.2 | 76.7 | 28.6 | 26.8 |

<sup>a</sup>The peak area for each allergen was expressed as the % peak area relative to the total peak area in the HPLC chromatogram and assigned 100% for light roast peanut flour. The % peak areas for each allergen in medium and dark roast peanut flour are shown relative to the peak area of the corresponding allergen in light roast peanut flour.
<sup>b</sup>Ara h 1 peak elutes as a shoulder in the front side of a large Ara h 3 peak. The Ara h 1 peak is not recognized by the HPLC peak integration software in the medium roast peanut flour due to extensive degradation of the major Ara h 3 peak. Therefore, the Ara h 1 peak area was estimated as roughly 30% of the peak area where normally Ara h 1 elutes.
**Abbreviation:** HPLC, high-performance liquid chromatography.

### 3.2 Relative potency

The relative potency data (allergen levels) for immunodominant peanut protein allergens varied by about 3-fold within each of the three allergen ELISA tests (**Figure 3**). Relative potency data among the screened peanut flour lots also show seasonal variability. Controlling potency to a tight fold range minimizes the potency variability between each dose-increase step in the multistep dose-escalation treatment. To ensure the consistency of PTAH potency, the range of variation in the allergens are tightly controlled by the peanut flour pre-selection process. The actual range of potency within selected peanut flour lots was even narrower than 3-fold. The presence and consistency of other allergens of lesser clinical importance (Ara h 3, Ara h 7, Ara h 8, Ara h 9, and Ara h 10 through Ara h 17) were also characterized in peanut flour lots.

**Figure 3.**
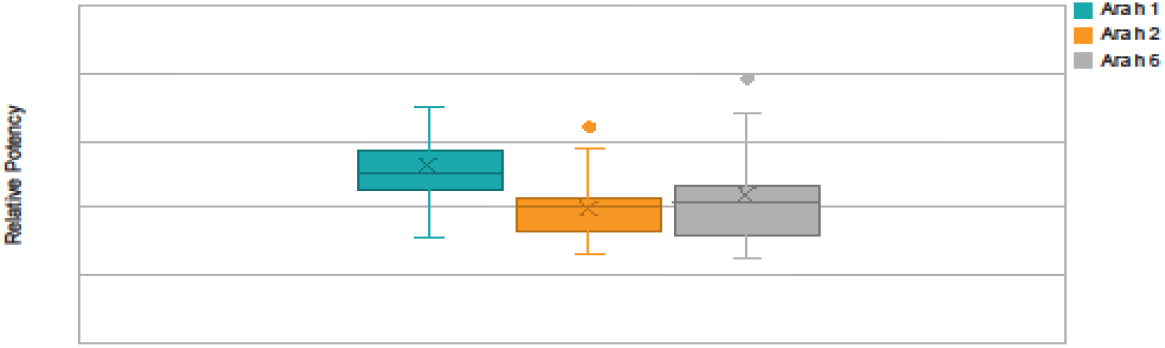
Relative Potency Ranges for Ara h 1, Ara h 2, and Ara h 6 of Selected Lots From 2018 to 2021. Whisker plot: The upper and lower whisker bars represent the upper and the lower extreme values. The upper and lower boundaries of the box and the horizontal line represent the upper and lower quartiles and the median. X is the mean. Single data point is an outlier.

### 3.3 Relative amounts of protein allergen

HPLC testing allows determination of whether allergens are present, their relative abundance, and if they are intact and not degraded within the peanut flour (ie, quality). Levels of allergens within a particular lot and lot-to-lot comparisons with the control standard are also determined by HPLC and ensure consistency. Based on the ranges and integrity of the allergens by HPLC, relatively narrow variation was observed in the relative amount of protein allergens (**Figure 4**).

**Figure 4.**
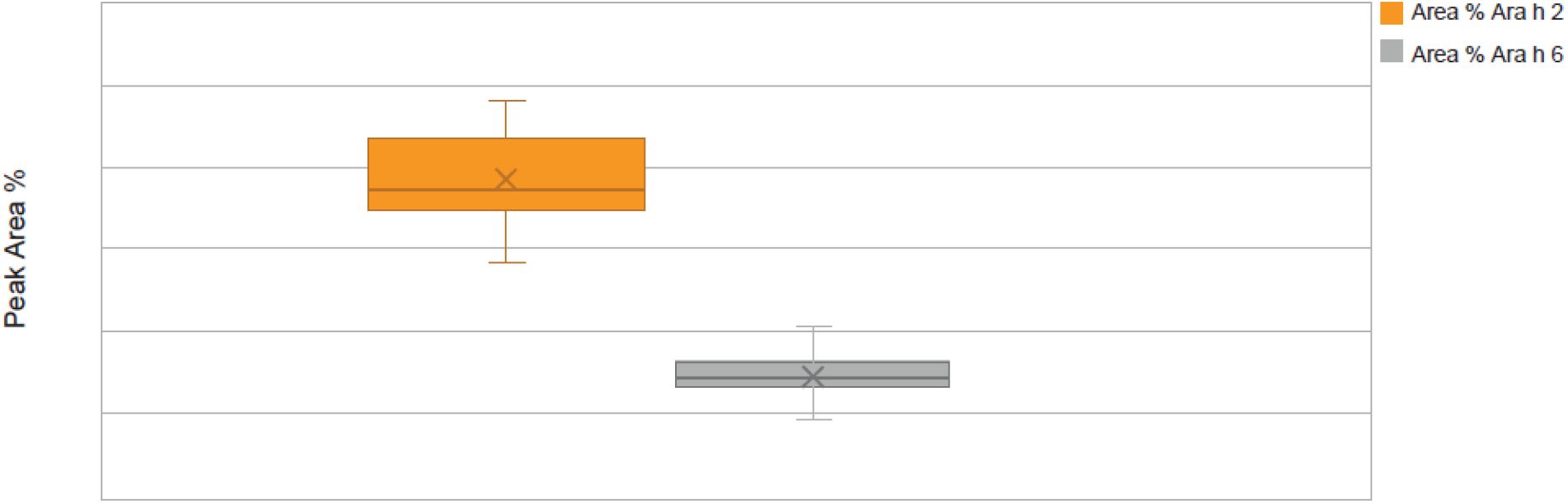
Allergens Ara h 2 and Ara h 6 Lot Peak Area Percentage Ranges by Protein Integrity From 2018 to 2021 in Source Material Screened Lots. Whisker plot: The upper and lower whisker bars represent the upper and the lower extreme values. The upper and lower boundaries of the box and the horizontal line represent the upper and lower quartiles and the median. X is the mean. Ara h 1 is not displayed due to a very small peak area percentage.

### 3.4 Aflatoxin levels

The proportion of the lots not meeting the aflatoxin limits has increased in recent years (2011 to 2020) (**Figure 5**). A significant proportion of commercial peanut flour lots did not meet aflatoxin total limits for PTAH drug substance use. Total aflatoxin content in commercial peanuts for human consumption is limited to 15 ppb according to the Code of Federal Regulations, Title 7, Part 996.11 (29). Peanut flour lots used in PTAH manufacturing contain substantially stricter limits of total aflatoxins and in particular, aflatoxin subspecies B_1_, to provide a safety margin and to conform to the quality standards required by the European Commission Regulation 1881/2006, Annex 2.1.3 (30). As shown in **Figure 5**, many of the lots surpass the manufacturing limit for aflatoxin contamination and are rejected as source material for PTAH.

**Figure 5.**
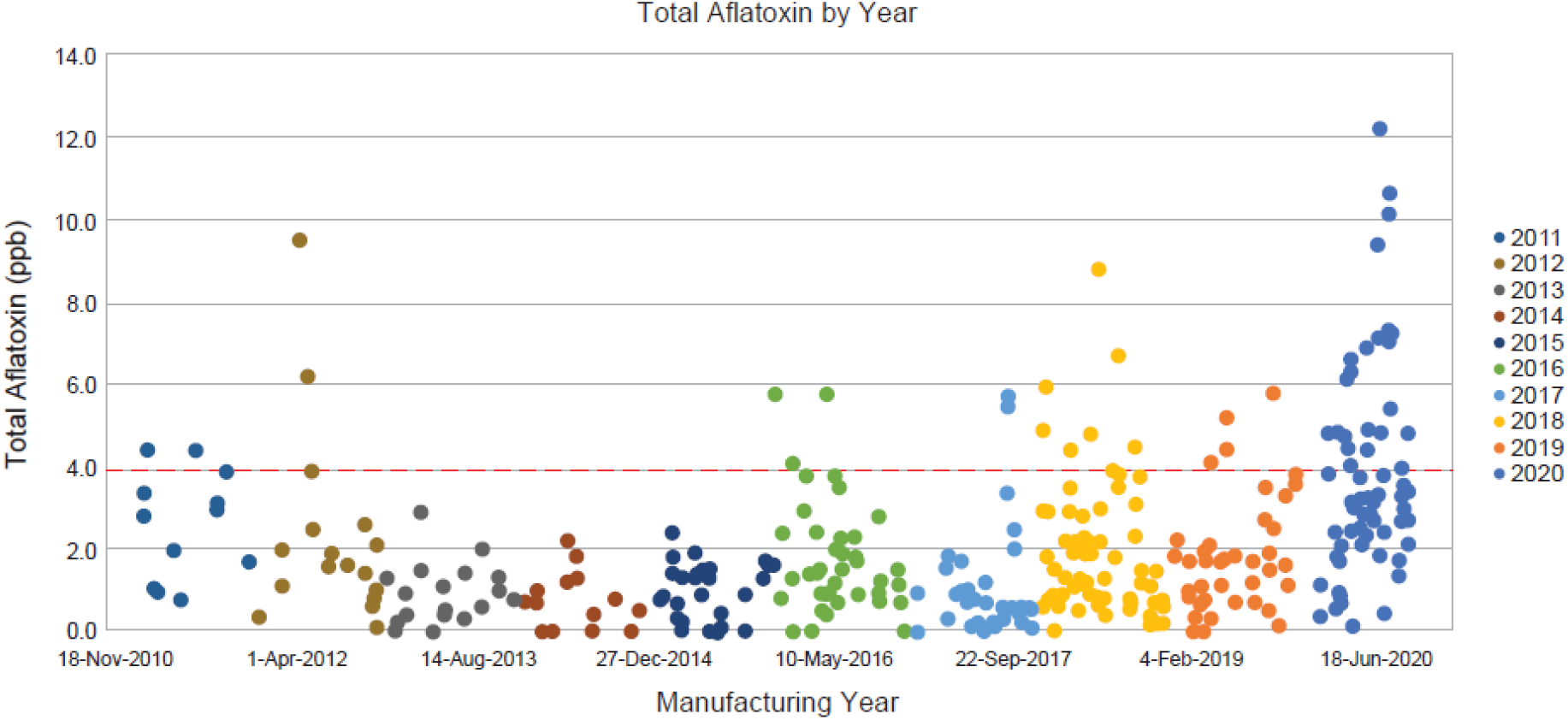
Total Aflatoxins in Commercial Peanut Flour Lots From 2010 to 2020. Peanut flour lots used in PTAH manufacturing are limited to no more than 4 ppb of total aflatoxins, shown by the red dashed line. GPTN would not manufacture peanut flour from peanut crop where the total aflatoxin exceeds 15 ppb. **Abbreviations:** GPTN, Golden Peanut and Tree Nuts; ppb, parts per billion; PTAH, peanut (*Arachis hypogaea*) allergen powder-dnfp.

### 3.5 Peanut flour lot rejection overall

Overall, in recent years, the majority (up to 60%) of peanut flour source material failed to meet screening selection acceptance criteria for additional drug substance testing (**Table 4**). The most common reasons for lot rejection included failure to meet relative potency acceptance criteria for one or more component allergens, as well as total aflatoxin level near or higher than the drug substance acceptability threshold. For aflatoxin, rejected lots often differed from acceptable limit by several fold.

**Table 4.** Historical Data for Peanut Flour Source Material Lots Screened and Rejected From Selection for Drug Substance.

| **Year** | **Number of Source Material Lots Selected for Screening** | **Number of Lots Rejected for Drug Substance Selection** | **Percentage of Screened Lots Rejected as Unsuitable for Drug Substance Testing** |
| --- | --- | --- | --- |
| 2018 | 14 | 8 | 57% |
| 2019 | 8 | 3 | 38% |
| 2020 | 7 | 4 | 57% |
| 2021 | 5 | 3 | 60% |

### 3.6 Correlation/Association with clinical outcomes

The specification range for relative potency by ELISA test for each allergen is approximately 3-fold, which is comparable to the limit of standardized venom or inhalant allergen products (8). The actual relative potency of clinical lots of PTAH was controlled well within the specification limits range. The likelihood of a patient receiving a dose level with a low potency lot followed by an up-dose level with a high potency lot is, therefore, lower than if the allergen had not been standardized and maintained in a specified range. This is especially important for up-dosing where incrementally higher doses are given over time. Control of potency minimizes the risk of a large variability in potency between dose levels, that is, if a PTAH lot at the lower specification limit is used to dose a patient at one dose level and another lot at the upper specification limit is used at the following dose-escalation step.

The PTAH dosing protocol involves stepwise dose increases in peanut protein content that range from 1.2-fold to 2.0-fold in magnitude, except for a 0.5-fold decrease in intended dose between 6 mg at the end of initial dose escalation and 3 mg at the start of the up-dosing phase of treatment. Potency analyses were performed on drug product capsule and sachet lots used in two PTAH phase 3 clinical trials. These assessments represented a total of 5246 participant up-dosing experiences in 520 patients (**Table 5**). For the up-dosing experiences, the frequency distribution of various ratio dose escalations due to both dose and potency in the two clinical studies was analyzed. When the measured potencies of drug product lots were applied to the intended dose increases, 95% of escalations ranged from 0.87 to 2.38, 1.00 to 2.50, and 1.00 to 2.25 for Ara h 1, Ara h 2, and Ara h 6, respectively.

**Table 5.** Summary of the Relative Potency Ranges and Potency Ratios of Capsule and Sachet Lots Used in ARC003 and ARC010 Clinical Studies.

| Clinical Study | Number of Drug Product Batches | Ara h 1 |  | Ara h 2 |  | Ara h 6 |  |
| --- | --- | --- | --- | --- | --- | --- | --- |
|  |  | Relative Potency Range | Ratio<sup>a</sup> | Relative Potency Range | Ratio<sup>a</sup> | Relative Potency Range | Ratio<sup>a</sup> |
| PALISADE (ARC003) | 8 | 0.68-1.59 | 2.34 | 0.72-1.34 | 1.86 | 0.63-1.26 | 2.00 |
| ARTEMIS (ARC010) | 11 | 0.68-1.25 | 1.83 | 0.93-1.34 | 1.44 | 0.75-1.26 | 1.68 |
| Total | 19 | 0.68-1.59 | 2.34 | 0.72-1.34 | 1.86 | 0.63-1.26 | 2.00 |

The data shown in the table are from a total of 5246 individual up-dosing events for 520 patients.
<sup>a</sup>Ratio is from highest to lowest potency.

The 5246-participant up-dosing events discussed above included close monitoring in a clinic; adverse events were reported in both PTAH-treated and placebo-treated patients. All but one of these events was graded as either mild or moderate severity. A single severe reaction occurred during an up-dosing visit with 200 mg (after previously taking a 160 mg daily dose). Potency analysis of the lot of drug product used during this visit revealed relative potencies of 1.14, 0.87, and 0.90 for Ara h 1, Ara h 2, and Ara h 6, respectively, which are near the center of the range of the potencies of the lots used. Based on this evaluation, the likelihood that potency variation accounted for the severity of the clinical reaction was considered low.

## 4 Discussion

In 1911, Leonard Noon published a report of allergen immunotherapy used for allergic rhinitis caused by grass pollen in the United Kingdom (31). The observation that administering incrementally increasing amounts of an allergen to an allergic person could lead to a “desensitized” state, resulting in symptom improvement, has led to immunotherapy strategies utilizing inhalant allergens, stinging insect venoms, and more recently, foods (32-34).

During the first half of the 20^th^ century, the standard of practice for subcutaneous immunotherapy evolved without regulatory guidance or the benefit of placebo-controlled trials to evaluate safety and efficacy (34, 35). Empiric and anecdotal application of immunotherapeutic principles to treat allergic diseases became widely accepted, but also unintentionally led to routine inclusion of some “allergens” with no efficacy (eg, body bee extract), and in some cases, involved potentially unsafe practices (eg, administration of subcutaneous allergen injections to patients with poorly controlled asthma or allowing routine home administration of subcutaneous immunotherapy injections) (36, 37). Safety of allergen immunotherapy might be considered 2-fold, involving safety of the drug product itself (ie, protection from harm due to variability in potency or contaminants/impurities) and clinical safety (ie, protection from harm due to biological/physiological effects of the drug when taken by an individual). The nature of drug safety in individuals with peanut allergy is likely heterogeneous; however, ensuring the drug product is high quality and consistent reduces concern that clinical safety is confounded by or due to hazards arising from the drug product.

Adherence to regulations for quality of allergen-specific immunotherapy in Europe and the US is required to obtain marketing approval or authorization (6, 7, 38, 39). Lot-to-lot consistency and shelf life stability (influenced by stability of individual drug components) are critical to ensure quality, therefore these regulations guide presence (within specific ranges and including justification for selection) of relevant allergens, consistency of protein content (within specific ranges), and limits on impurities (14, 38). While the preference toward products with proven quality, safety, and efficacy has been demonstrated worldwide over the last 20 years and requirements have been implemented to distinguish allergen drug products for immunotherapy for food allergy from non-industrial preparations of allergen immunotherapy directly from food sources that are less controlled and standardized, challenges do arise for analytical characterization of food allergens and correlations between biological potency and protein content in the assessment of quality for allergen immunotherapies (2, 39). Additionally, regulatory guidance appears to be more specific for aeroallergens and insect venom allergies than for food allergen immunotherapy products (39). Use of allergen immunotherapy may be limited by the availability of high-quality, standardized drug products with proven efficacy and safety, as recommended by professional organizations (i.e., EAACI, AAAAI, and ACAAI) (8, 13).

Sourcing peanut flour for OIT treatment from GPTN, a food-grade peanut manufacturer, is the starting point for drug substance manufacturing. This peanut flour source material had already undergone substantial analysis and met important quality criteria, yet less than 50% of GPTN lots were suitable for use as drug substance in the PTAH GMP manufacturing process. A fundamental requirement for an approved drug is thorough confirmation of drug identity, quality, and safety through all phases of product manufacture (ie, raw materials, drug substance, in-process, to final drug product), and this is facilitated by adhering to GMP (40). These processes ensure that each packaged dose of drug product meets strict criteria for many attributes throughout its shelf life, including physical, chemical, and immunological properties. However, failure to meet acceptance criteria for potency— relative to an in-house reference standard—for each of the immunodominant allergens, Ara h 1, Ara h 2, and Ara h 6, most often accounted for rejection of peanut flour source material, followed by failure to meet acceptance criteria for aflatoxin contamination. In other words, to meet acceptance criteria, peanut flour lots must have appropriate levels of immunodominant allergens and lower levels of aflatoxin contamination.

A drug substance reference standard is rigorously qualified using all the tests on the drug substance manufacturer’s COA, as well as highly characterized using additional analytical methods (US Pharmacopeia or European Pharmacopoeia) that are a necessary activity in the process of drug standardization. As mentioned previously, when preparing allergen for use as subcutaneous immunotherapy treatment, practice guidelines and regulatory authorities suggest choosing standardized allergens when available because of the safety and efficacy advantages of limiting potency variation (8, 12). No such guidelines currently exist for allergens used as food OIT, but the regulatory pathway for the commercial development of an OIT for food allergy has been clarified. Food, when used for medicinal use as a treatment of a food allergy, is considered by regulatory authorities to be a biologic drug, which is regulated in the US by the FDA’s Center for Biologics Evaluation and Research (6, 33). The European Pharmacopoeia specifies quality requirements including processes and methods on manufacturing and analysis of medicinal products (38). As such, allergen standardization requires a reference standard(s) as well as a thorough confirmation of its identity, quality, potency, and safety through all phases of product development (12). The reference standard is used to ensure lot-to-lot consistency of allergenic potency. It should be noted that PTAH contains all relevant peanut allergens (as well as the natural mixture of proteins present in peanut, Ara h 1 through Ara h 17) (24) and that each lot of PTAH drug substance has met acceptance criteria for relative potency of the immunodominant and the most clinically relevant component allergens Ara h 1, Ara h 2, and Ara h 6 using ELISA; the presence, identity, intact form, and relative levels of these component allergens also met acceptance criteria for consistency relative to both each other and to other lots using reverse-phase HPLC (27, 28).

Extensive variability in allergen potency (e.g., if uncontrolled food stuff containing peanut is used as OIT) is particularly important to consider during OIT up-dosing visits when the allergen dose is sometimes intentionally stepped up in 2-fold or more increments. Such variation would present a risk that a lower than intended potency of a component allergen (e.g., Ara h 1) would be up-dosed to a higher than intended potency, even when the weight of the peanut flour is appropriate for the intended dose (8). The relevance of this concern is illustrated by the fact that the natural variation in peanut component allergen potency is substantial, even in high-quality, commercially available peanut flours or other peanut containing products. For example, in one study, the ratio of Ara h 2 to Ara h 1 in a given weight of peanut flour varied more than 40-fold (0.56 to 23.30), depending on the type and source of the peanut flour (9). With a standardized allergen and regulatory body–approved medicine, this kind of potency variation is avoided, as evidenced by the consistent relative potencies of Ara h 1, Ara h 2, and Ara h 6 in our analysis of the PTAH lots used in the PTAH phase 3 clinical studies.

## 5 Conclusions

Extensive variability in allergen potency is, in part, due to combined seasonal and peanut flour manufacturing process variations and is an important consideration for a therapy used in OIT. This is particularly critical during PTAH up-dosing (dose escalation), as substantial variability may result in lack of efficacy or trigger an adverse allergic reaction. Over the years, the source material for peanut flour has shown extensive variation in relative potency, protein component content, and aflatoxin levels. Rigorous GMP manufacturing process and testing controls have been implemented based on EU and US regulatory requirements to ensure product safety, potency, quality, and safety, at both initial manufacture, as well as through the end of shelf life of PTAH drug product. The rigor of PTAH’s manufacturing process ensures product consistency between lots and reduces the risk of unintended clinical safety and efficacy outcomes.

## Data Availability

All data produced in the present study are available upon reasonable request to the authors

## Conflict of interest

**Stephanie A. Leonard** reports being a member of the International FPIES Association medical advisory board.

**Yasushi Ogawa** is an employee of Aimmune Therapeutics, a Nestlé Health Science company.

**Paul T. Jedrzejewski** is an employee of Aimmune Therapeutics, a Nestlé Health Science company.

**Soheila J. Maleki** has no disclosures to report.

**Martin D. Chapman** reports an R01 research grant on the structural biology of allergens from NIH NIAID. In addition, they report honorarium for molecular allergology symposium from Johns Hopkins University and is a co-owner and shareholder of Indoor Biotechnologies.

**Stephen A. Tilles** is an employee of Aimmune Therapeutics, a Nestlé Health Science company.

**George Du Toit** reports research grants to their institution and advisory board fees from Aimmune Therapeutics, a Nestlé Health Science company.

**S. Shahzad Mustafa** reports honoraria for Aimmune program.

**Brian P. Vickery** reports advisory board/consultant for Aimmune Therapeutics, AllerGenis, FARE, Reacta; site investigator for Aimmune Therapeutics, DBV, Genentech, Regeneron; and research grants from FARE and NIAID.

## Author contributions

All authors agree to be accountable for all aspects of the work.

**S.A.L**. contributed to analysis/interpretation of data; part of the manuscript writing team and approves the version for publication.

**Y.O**. contributed to data collection/analysis/interpretation of data; part of the manuscript writing team and approves the version for publication.

**P.T.J**. contributed to data collection/analysis/interpretation of data; part of the manuscript writing team and approves the version for publication.

**S.J.M**. contributed to analysis/interpretation of data; part of the manuscript writing team and approves the version for publication.

**M.D.C**. contributed to analysis/interpretation of data; part of the manuscript writing team and approves the version for publication.

**S.A.T**. contributed to analysis/interpretation of data; part of the manuscript writing team and approves the version for publication.

**G.D.T**. contributed to analysis/interpretation of data; part of the manuscript writing team and approves the version for publication.

**S.S.M**. contributed to analysis/interpretation of data; part of the manuscript writing team and approves the version for publication.

**B.P.V**. contributed to analysis/interpretation of data; part of the manuscript writing team and approves the version for publication.

## Funding

This study was sponsored by Aimmune Therapeutics, a Nestlé Health Science company.

## Data sharing statement

The data supporting the findings of this study are available within the article its supplementary materials. Clarifying requests can be made to the corresponding author.

## Acknowledgments

The authors wish to acknowledge the contributions of Andrea Vereda and Anne-Marie Irani for review and helpful comments during the development of this manuscript. Aimmune Therapeutics, a Nestlé Health Science company, provided financial support for this manuscript. Writing and editorial assistance for this manuscript was provided by Stephanie Phan, PharmD, and Cheryl Casterline, MA (Peloton Advantage, LLC, and OPEN Health Company, Parsippany, NJ).

## Notes

### Funding Statement

This study was sponsored by Aimmune Therapeutics, a Nestle Health Science company.

### Author Declarations

The data from clinical trials NCT03201003 and NCT02635776 (available at clinicaltrials.gov) have previously been published here: PALISADE Group of Clinical Investigators, Vickery BP, Vereda A, Casale TB, Beyer K, du Toit G, et al. Ar101 oral immunotherapy for peanut allergy. N Engl J Med (2018) 379(21):1991-2001. Epub 2018/11/20. doi: 10.1056/NEJMoa1812856. Hourihane JOB, Beyer K, Abbas A, Fernandez-Rivas M, Turner PJ, Blumchen K, et al. Efficacy and safety of oral immunotherapy with ar101 in european children with a peanut allergy (artemis): A multicentre, double-blind, randomised, placebo-controlled phase 3 trial. Lancet Child Adolesc Health (2020) 4(10):728-39. Epub 2020/07/24. doi: 10.1016/S2352-4642(20)30234-0.

