## Supplementary Information for "Manufacturing Processes of Peanut (*Arachis hypogaea*) Allergen Powder-dnfp"

### *Supplementary Material*

#### 1 Importance of standardization

Manufacturing of allergen extracts may involve roasting, grinding, defatting, extraction, clarification, and sterilization (10). Each of these processes may introduce variabilities that contribute to the lot-to-lot differences of the final product (10). Standardization is a process that allows for potencies of similar products from different manufacturers and manufactured lots to be compared (10).

Standardization of a drug through confirmation of its identity, quality, and purity through all phases of product development is critical because all of these factors contribute directly or indirectly to the safety, effectiveness, and acceptability of a product (2, 33). The lack of agreed-upon or evidence-based measures or guidelines for establishing quality can lead to health problems.

Because there is a lack of clinician-generated peanut preparations used for allergen immunotherapy and a lack of standardization in food products, individuals with peanut allergy may face barriers obtaining immunotherapies with precise and consistent allergen profiles. Because peanut flour is naturally derived, its potency varies. Drug identity, quality, and purity play a role in contributing to the safety, efficacy, and acceptability of a drug.

#### 2 Peanut flour source material and additional detail on roasting and defatting

The peanut flour source material for PTAH is manufactured exclusively from jumbo Runner kernels. Runner-type peanuts are the primary cultivar grown in the Southeastern US, accounting for approximately 85% of total US peanut production. Raw, shelled jumbo Runner peanuts are roasted and pressed in an all-natural oil extracting process (no solvents used) to deliver 12% fat level ingredients. Roasting significantly changes potency and affects protein integrity of extracted peanut allergens, thus impacting peanut flour quality (9, 10). Greater heat exposure is associated with more degradation of the peanut allergens, as observed in the high-performance liquid chromatography (HPLC) profiles of medium and dark roasted peanut flours.

Roasting of raw shelled peanuts consists of loading peanuts in a feed hopper to be moved by a conveyer belt into a continuous forced-air convection dry roaster. Peanuts also pass through a metal detector prior to roasting and a magnet is used to ensure removal of metallic contaminants. The conveyer belt controls roasting time within a range of speeds and temperatures in various zones. These limits have been demonstrated to provide a substantial reduction in *Salmonella* species through regular validation of the roasting process. Roasted peanuts are processed through the blancher (de-skinner), where the peanut skins are separated from the peanut kernels by mechanical action and the skins are then removed by aspiration (ie, air conveyance). Roasted de-skinned peanut kernels are ground into a paste that is pumped into a hydraulic press to reduce the fat content to approximately 12%. Fat content of the finished source material is measured during processing using a near-infrared, in-process method, and the press dwell time is adjusted as needed to ensure that the fat content of the partially defatted peanut flour (source material) remains within the established specification range.

Subsequently, partially defatted peanut press cake is discharged from the hydraulic press and passed through a rasp-type mill to convert it to peanut flour. Peanut flour passes through a series of rare-earth magnets and then through a metal detector to ensure removal of any metal contaminants.

#### 3 Pesticides

Since pesticides that are approved for use on peanuts as food materials may be used by peanut growers during the growing, harvesting, and storage of farmers' stock jumbo Runner peanuts prior to the shelling process, residual pesticides from application and use of these chemicals are controlled in the allergen source material by testing performed at a contract test laboratory that conducts pesticide characterization testing on food products (in compliance with provisions of International Organization for Standardization [ISO] 17025, General Requirements for the Competence of Testing and Calibration Laboratories). A list of residual pesticides and the acceptance limit for each pesticide have been established for the testing performed on the allergen source material peanut flour.

#### 4 Additional details on primary and secondary reference standards

Potency of the secondary reference standard is normalized to the potency of the primary reference standard, and a correction factor is calculated. The correction factor is applied to the potency of test samples when a sample is tested against the secondary reference standard to normalize the test results relative to the potency of the primary reference standard. The primary reference standard is also used to qualify a new lot of secondary reference standard to minimize drift in the potency of secondary reference standards by establishing a correction factor for each lot of secondary reference standard. Stability of the primary and secondary reference standards is monitored by testing annually. This process ensures the reference standard in use has remained stable through its assigned expiry. The reference standard is used in relative potency ELISA to test samples against the reference standard, and the potency of the test samples is normalized to the potency of the reference standard and reported as a relative potency. The reference standard is used as the reference peak retention time for peanut allergens in protein integrity HPLC profile of peanut flour samples. Peak retention times of the allergen peaks in the sample must agree with the retention times of the peaks in the reference standard chromatogram. The overall chromatographic profile of the sample must also match the chromatographic profile of the reference standard. The reference standard is also used in assay, which measures and reports total protein content of the peanut flour sample. The reference standard serves as a system suitability control for total protein content in assay.
